# Prognostic Implications of Right Ventricular Dysfunction in Degenerative Mitral Regurgitation

**DOI:** 10.1101/2023.11.10.23298404

**Authors:** Yohann Bohbot, Benjamin Essayagh, Giovanni Benfari, Jeroen J. Bax, Thierry Le Tourneau, Yan Topilsky, Clemence Antoine, Dan Rusinaru, Francesco Grigioni, Nina Ajmone Marsan, Aniek van Wijngaarden, Aviram Hochstadt, Jean Christian Roussel, Prabin Thapa, Hector I Michelena, Maurice Enriquez-Sarano, Christophe Tribouilloy

## Abstract

**Background:** The prevalence and impact of right ventricular dysfunction (RVD) in degenerative mitral regurgitation (DMR) is unknown. We aimed to determine whether RVD assessed by echocardiography in routine clinical practice is independently associated with survival in patients with DMR.

**Methods:** We used data from the Mitral Regurgitation International DAtabase-Quantitative (MIDA-Q) which included patients with isolated DMR due to mitral valve prolapse from January 2003 to January 2020 from 5 tertiary centers across North America, Europe, and the Middle East. A cohort of 4,379 (mean age: 67 years, 64.6% males, follow-up:5.3[3.3-8.2]years) consecutive patients with significant (≥moderate) DMR was included and long-term survival was analyzed.

**Results:** RVD, identified in 584(13.3%) patients, was associated with reduced 10-year survival (49±3% *vs.* 67±1%; *p<0.001*), overall and in all subgroups of patients, even after comprehensive adjustment including left ventricular dilatation and dysfunction, DMR severity, pulmonary pressures and surgery (adjusted HR[95%CI]=1.55[1.31-1.84]; *p<0.001*). This excess mortality was observed under medical management (adjusted HR[95%CI]=1.39[1.12-1.72]; *p=0.003*) and after surgical correction of mitral regurgitation (adjusted HR[95%CI]=1.56[1.08-2.27]; *p=0.019*). Patients with RVD undergoing surgery within 3 months of diagnosis experienced a better 10-year survival (73±4% vs. 38±4%; *p<0.001*), even after adjustment (adjusted HR[95%CI]=0.53[0.35-0.81]; *p=0.003)* despite an increase of 1-month post-operative mortality (5% vs 2.2% for patients without RVD; *p<0.001*).

**Conclusions:** RVD is observed in 13.3% of significant DMR and exhibits a powerful and independent association with excess mortality partially attenuated by mitral surgery. Therefore, assessment of right ventricular systolic function should be included in routine DMR evaluation and in the clinical decision-making process.

## INTRODUCTION

Right ventricular (RV) systolic dysfunction is a major determinant of survival in most cardiovascular diseases as heart failure(1), pulmonary hypertension (PH)(2) and myocardial infarction.(3) Conversely, in the field of left-sided valvular heart disease (VHD), considerable attention has been paid to left ventricular dysfunction, but RV function has been somewhat neglected. One possible explanation is that RV dysfunction (RVD) is often perceived as secondary to PH which is a strong and recognized prognostic factor in left-sided VHD(4–7), particularly in degenerative mitral regurgitation (DMR).(5,6) Indeed, the chronic increase in RV afterload induced by PH can lead to progressive RV failure with a negative correlation between RV function and pulmonary artery pressure.(8) Few studies of limited size have suggested that RVD assessed by radionuclide cineangiography is associated with poor outcome in patients with DMR(9,10) but without addressing the link with PH. In a study of 208 patients published 10 years ago, Le Tourneau et al. reported that impaired RV function assessed by radionuclide angiography is common, depends weakly on pulmonary artery systolic pressure and conveys important prognostic information in patients with chronic DMR.(11) Nevertheless, to date, assessment of RV function is not integrated into the clinical decision-making process in DMR(12,13) and no large-scale data is available regarding the impact of RV function assessed by echocardiography in these patients.

In this context, we sought to evaluate the prevalence of RVD diagnosed by transthoracic echocardiography in routine clinical practice using a large multicenter cohort of patients with ≥ moderate mitral regurgitation (MR) of degenerative origin and to assess its independent impact on survival, under medical management and after surgical correction of MR.

## METHODS

### Study design

We used data from the Mitral Regurgitation International DAtabase-Quantitative (MIDA-Q) registry, corresponding to the merging of electronic databases of patients with isolated DMR quantified prospectively in routine clinical practice of tertiary centers in Europe (University of Amiens, Amiens, France; University of Nantes, Nantes, France; Leiden University Medical Center, Leiden, the Netherlands), North America (Mayo Clinic, Rochester, MN, USA), and the Middle East (Tel Aviv Medical Center, Tel Aviv, Israel). Patient recruitment ranged from January 2003 to January 2020, depending on each center’s database.(14,15)

Consecutive adult patients (aged 18 years or older) with a diagnosis of mitral valve prolapse with or without a flail leaflet and a comprehensive clinical and echocardiographic evaluation at diagnosis, were eligible for inclusion. We excluded patients: (i) who declined research authorization, (ii) without quantification of MR, (iii) with functional or rheumatic MR, ≥ moderate aortic valve disease or mitral stenosis, congenital heart disease, active endocarditis, dilated/hypertrophic/restrictive cardiomyopathies and (iv) with a history of valvular surgery. The present analysis was based on a study of patients with significant (≥moderate) DMR. The study was approved by the Institutional Review Board of each center and conducted in accordance with institutional guidelines, national legal requirements, and the revised Declaration of Helsinki.

### Echocardiography

All patients underwent comprehensive Doppler-echocardiographic assessment using commercially available ultrasonography systems. All echocardiographic studies were analyzed by experienced investigators from each center. Left ventricular (LV) dimensions were assessed from parasternal long-axis views by 2-dimensional-guided M-mode, using the leading-edge methodology at end-diastole and end-systole. Left ventricular ejection fraction (LVEF) was calculated using Simpson’s biplane method (16). Left atrial volume index (LAVI) was estimated using the biplane method from apical 4-and 2-chamber views and indexed for body surface area (BSA).(16) MR was graded as moderate or severe according to current recommendations using a multiparametric approach, which included estimation of effective regurgitant orifice area (EROA) and regurgitant volume of MR using the PISA technique.(17,18) Pulmonary artery systolic pressure (sPAP) was measured by applying the modified Bernoulli equation using the tricuspid regurgitation peak jet velocity and adding estimated right atrial pressure according to the inferior vena cava diameter and its respiratory variation. RV size and function was assessed as recommended per current guidelines using comprehensive imaging with multiple windows examining all segments of the complex right ventricular anatomy and with the presence of RV dysfunction diagnosed based on a combination of qualitative and quantitative measures, including assessment of fractional area change, tissue-Doppler S’ and tricuspid annular plane systolic excursion (TAPSE). (16) The final diagnosis of RV dysfunction by the cardiologist responsible for the Doppler-Echocardiographic examination final report was based on the integration of all information available in a categorical classification as per guidelines. (16)

### Follow-up and Endpoints

Median follow-up was 5.3 (interquartile range [IQR]: 3.3 to 8.3) years. Events were ascertained by direct patient, family, or referent physician contact and by using institutional, private (Accurint in the United States), or public (social security mortality database or local equivalent) databases of vital status.(14) The primary end point was overall survival after diagnosis, with medical and surgical treatment. Secondary endpoints were survival under medical management in the overall population with censoring at mitral surgery (for patients operated on during follow-up), and post-operative survival in patients who underwent mitral surgery.

### Statistical analysis

The study population was divided in 2 groups according to RV function. Categorical variables were reported as percentages and counts and continuous variables were expressed as mean value±1 standard deviation or median (IQR). The relationship between baseline variables and the two groups was explored using 1-way ANOVA tests (for normally distributed continuous variables) or Kruskal-Wallis tests (for non-normally distributed continuous variables) and Pearson’s χ² statistic or Fisher’s exact test for categorical variables. Factors associated with RVD were identified using multivariable logistic regression analysis. Estimated survival rates± standard error were estimated according to the Kaplan Meier method and compared with two-sided log-rank tests. Multivariable analyses of all-cause mortality were performed using Cox proportional hazards models adjusted for age, sex and recognized prognostic factors in DMR including atrial fibrillation (AF)(14,15,19), presence of symptoms, LV end systolic diameter (LVESD)≥ 40mm (14,20), LVEF≤ 60% (14,21), LAVI ≥ 60 ml/m²(14,15,22), sPAP≥ 50 mm Hg(5,14,15) and MR severity. Subgroup analyses were performed to determine the homogeneity of the association between RVD and mortality. The effect of RVD on the overall mortality risk was first estimated in each subgroup using a Cox univariate model and then formally tested for first-order interactions in Cox models by entering interaction terms separately for each subgroup. The limit of statistical significance was p < 0.05. All tests were 2-tailed. SPSS version 26.0 software (IBM, Armonk, New York) was used for statistical analysis.

## RESULTS

### Baseline characteristics

Among 10, 910 patients diagnosed between 2003 and 2020 with isolated DMR, 2723 were excluded because of the lack of MR severity quantification. Among the 8187 remaining patients, 4,379 (mean age 67 years, 64.6% males) had significant (≥ moderate) MR and available data on RV function. Among them, 2332 (53%) were from North America (United States) and 2047 patients (47%) were from Europe/Middle East. A bileaflet prolapse was observed in 2116 (48.2%), a posterior prolapse in 1805 (41.2%) and a flail leaflet in 1347 (32.7%) patients. Mean EROA was 39 ± 21 mm², mean LVESD was 35 ± 6 mm and mean LVEF was 63 ± 8% (***Table 1***).

**Table 1:** Baseline characteristics according to the presence of right ventricular dysfunction.

| Characteristics | Overall<br>n=4379 | Absence of RV<br>dysfunction<br>n=3795 | Presence of RV<br>dysfunction<br>n=584 | P-Value |
| --- | --- | --- | --- | --- |
| **Clinical characteristics** |  |  |  |  |
| Age, years | 67 ± 14 | 66 ± 14 | 74 ± 13 | <0.001 |
| Female gender (% , n) | 35.4 (1548) | 35.6 (1350) | 33.9 (198) | 0.43 |
| Body mass index, kg/m<sup>2</sup> | 25.5 ± 4.6 | 25.5 ± 4.5 | 25.6 ± 4.7 | 0.53 |
| Hypertension (% , n) | 33.1 (1450) | 33.1 (1257) | 33.0 (193) | 0.97 |
| Diabetes mellitus (% , n) | 7.2 (317) | 6.5 (246) | 12.2 (71) | <0.001 |
| Coronary artery disease (% , n) | 21.1 (925) | 20.3 (771) | 26.4 (154) | **0.001** |
| EuroSCORE II, % | 1.3 ± 1.1 | 1.2 ± 1.0 | 1.8 ± 1.5 | **<0.001** |
| Symptoms (% , n) | 54.9 (2369) | 53.0 (1983) | 67.1 (386) | **<0.001** |
| Atrial fibrillation (% , n) | 26.4 (1158) | 24.1 (914) | 41.8 (244) | **<0.001** |
| **<u>Echocardiographic characteristics</u>** |  |  |  |  |
| LV end-diastolic diameter, mm | 55 ± 7 | 56 ± 7 | 56 ± 8 | 0.77 |
| Indexed LV end-diastolic diameter, mm/m<sup>2</sup> | 30 ± 4 | 30 ± 4 | 31 ± 5 | 0.16 |
| LV end-systolic diameter, mm | 35 ± 6 | 35 ± 6 | 37 ± 8 | **<0.001** |
| Indexed LV end-systolic diameter, mm/m<sup>2</sup> | 19 ± 4 | 19 ± 3 | 20 ± 5 | **<0.001** |
| LV ejection fraction, % | 63 ± 8 | 64 ± 8 | 59 ± 11 | **<0.001** |
| LA volume index, mL/m<sup>2</sup> | 62 ± 27 | 60 ± 26 | 72 ± 32 | **<0.001** |
| EROA, mm<sup>2</sup> | 39 ± 21 | 39 ± 20 | 44 ± 23 | **<0.001** |
| MR regurgitant volume, ml | 60 ± 29 | 59 ± 29 | 65 ± 33 | **<0.001** |
| Bileaflet prolapse, (% , n) | 48.3 (2116) | 47.8 (1814) | 51.7 (302) | 0.078 |
| Posterior prolapse, (% , n) | 41.2 (1805) | 42.1 (1596) | 35.8 (209) | **0.004** |
| Flail Leaflet, (% , n) | 32.7 (1347) | 32.9 (1185) | 31.0 (162) | 0.40 |
| ≥Moderate to severe TR | 17.3 (744/4301) | 14.4 (535/3720) | 36.0 (209/581) | **<0.001** |
| TAPSE (mm)* | 22 ± 5 | 23 ± 4 | 14 ± 2 |  |
| PA systolic pressure, mmHg** | 40 ± 15 | 38 ± 13 | 52 ± 18 | **<0.001** |

Continuous variables are expressed as mean ±1 standard deviation and categorical variables are expressed as percentages and numbers.
Abbreviations: COPD: chronic obstructive pulmonary disease; EROA: effective regurgitant orifice; LA: left atrial; LV: left ventricular; NYHA= New York Heart Association; PA: pulmonary artery; TAPSE: tricuspid annular plane systolic excursion; TR: tricuspid regurgitation.
\* Available in 2186 patients, \*\*Available in 4168 patients

Patients with RVD (n=584; 13.3%) were older, more often diagnosed with diabetes mellitus and coronary artery disease, had higher EuroSCORE II and presented more frequently with symptoms and AF (all *p<0.001*). They had more severe MR with larger EROA and regurgitant volume, resulting in greater LVESD, LAVI and sPAP and lower LVEF (50.7% had a LVEF ≤60%) than patients with normal RV function (all *p<0.001*) **(*Table 1***).

On multivariable logistic regression, sPAP≥ 50 mm Hg (Adjusted OR[95%CI]=4.21[3.36-5.28]) and LVEF≤ 60% (Adjusted OR[95%CI]=1.92[1.54-2.40]) were the two factors most strongly associated with RVD, but age (Adjusted OR[95%CI]=1.04[1.03-1.05] per year increase), AF (Adjusted OR[95%CI]=1.32[1.05-1.66]), EROA≥ 40mm² (Adjusted OR[95%CI]=1.56[1.24-1.97]), and LVESD≥ 40mm (Adjusted OR[95%CI]=1.74[1.36-2.22]) were also independently associated with RVD.

### Long-term outcome impact of RV dysfunction

#### Outcome in the overall population

One thousand and forty (23.7%) deaths were recorded during follow-up. Ten-year estimated survival was 49±3% for patients with RVD *vs*. 67±1% for patients with normal RV function (Log Rank *p<0.001*) (***Figure 1A***).

**Figure 1:**
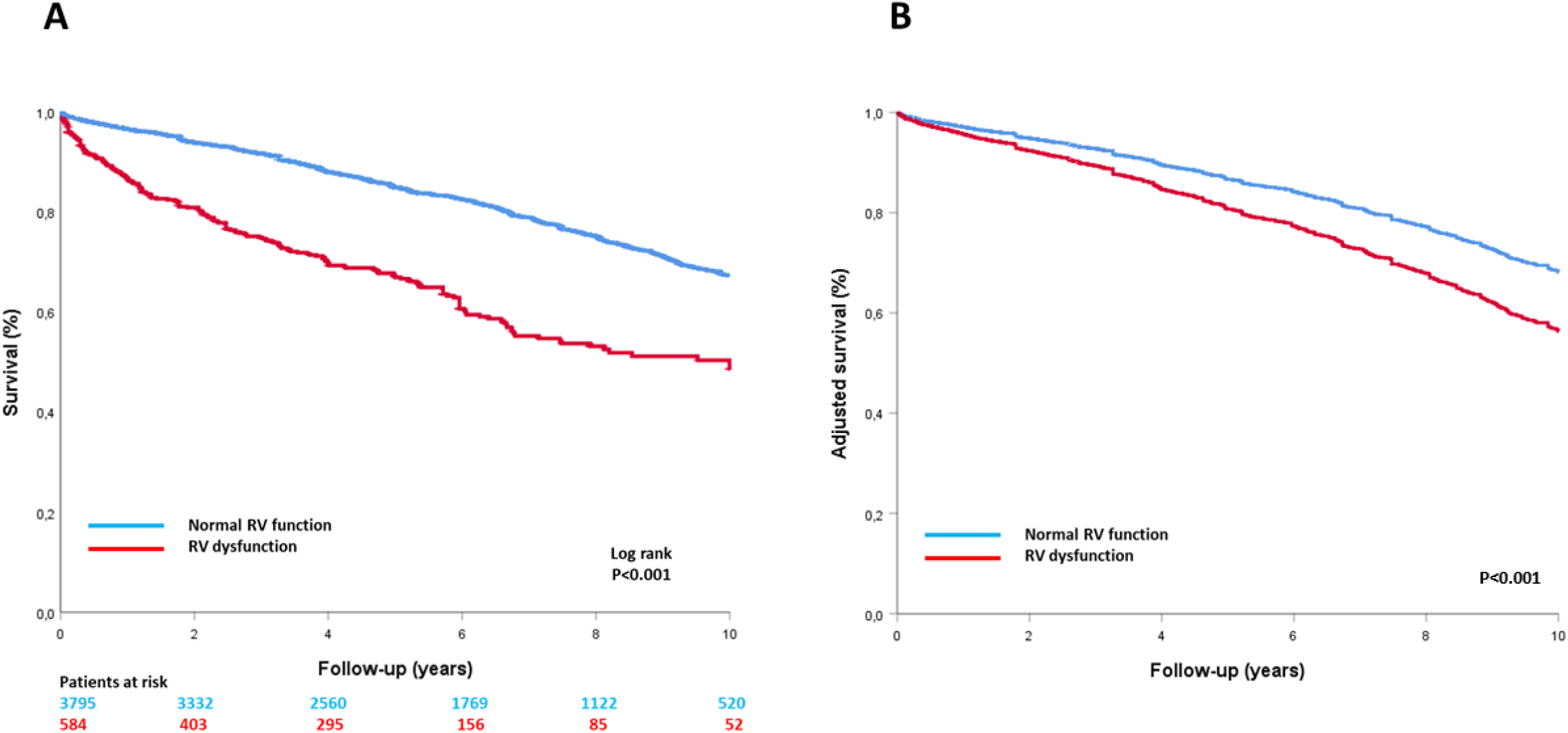
**Survival curves in the overall population.** Kaplan-Meier (A) and Cox adjusted (B) 10-year survival curves according to right ventricular function in the overall population. *RV: right ventricular*.

On Cox multivariable analysis, after adjustment for age, sex, presence of symptoms, AF, LVESD≥ 40mm, LVEF≤ 60%, LAVI ≥ 60 ml/m², sPAP≥ 50 mm Hg and MR severity, the impact of RVD on mortality was partially attenuated but persisted (adjusted HR [95%CI] =1.41 [1.18-1.68]; *p<0.001*) (***Table 2***, ***Figure 1B***). Results remained unchanged after further adjustment for mitral valve surgery treated as a time-dependent variable (adjusted HR [95%CI] =1.55 [1.31-1.84]; *p<0.001*). RVD remained independently associated with a poor survival in patients with sPAP<50 mmHg (adjusted HR [95%CI] =1.50 [1.12-1.92]; *p=0.001*) or ≥ 50 mmHg (adjusted HR [95%CI] =1.61 [1.24-2.08]; *p<0.001*);

**Table 2:** Univariable and multivariable hazard ratio for mortality for right ventricular dysfunction.

| Model | Overall population |  |
| --- | --- | --- |
|  | HR (CI[95%]) for RVD | P-Value |
| Univariable | 2.45 (2.12-2.85) | <0.001 |
| Adjusted for age ≥ 65 years, male sex, atrial fibrillation, symptoms and severe vs. moderate mitral regurgitation | 1.95 (1.68-2.28) | <0.001 |
| Further adjustment for LVESD ≥ 40 mm, LVEF ≤ 60%, LAVI ≥ 60 ml/m<sup>2</sup> and sPAP ≥ 50 mm Hg | 1.41 (1.18-1.68) | <0.001 |
| Further adjustment for mitral valve surgery (time-dependent) | 1.55 [1.31-1.84] | <0.001 |

Abbreviations: LAVI: left atria volume index; LVESD: left ventricular end-systolic diameter; LVEF: left ventricular ejection fraction; RVD: right ventricular dysfunction sPAP: systolic pulmonary artery pressure.

When the population was stratified according to stored TAPSE ≥ or < 17 mm (available in 2186 patients), the results in term of outcome was not different from the categorical RV function classification with an estimated 10-year survival of 70±2% for patients with TAPSE≥17mm *vs.* 46±4% for patients with TAPSE<17mm (*p<0.001*) and an adjusted hazard-ratio of 1.35[1.04-1.76]; p=0.024.

To verify the association of RVD with mortality in different population subsets, forest plot analysis was performed. Hazard ratios for mortality associated with RVD are presented in ***Figure 2*** for multiple subgroups based on clinical and echocardiographic variables. The association between RVD and risk of death was consistent in subgroups of patients with no interactions between RVD and any of the subgroups (All *p for interaction ≥ 0.16*) (***Figure 2***).

**Figure 2:**
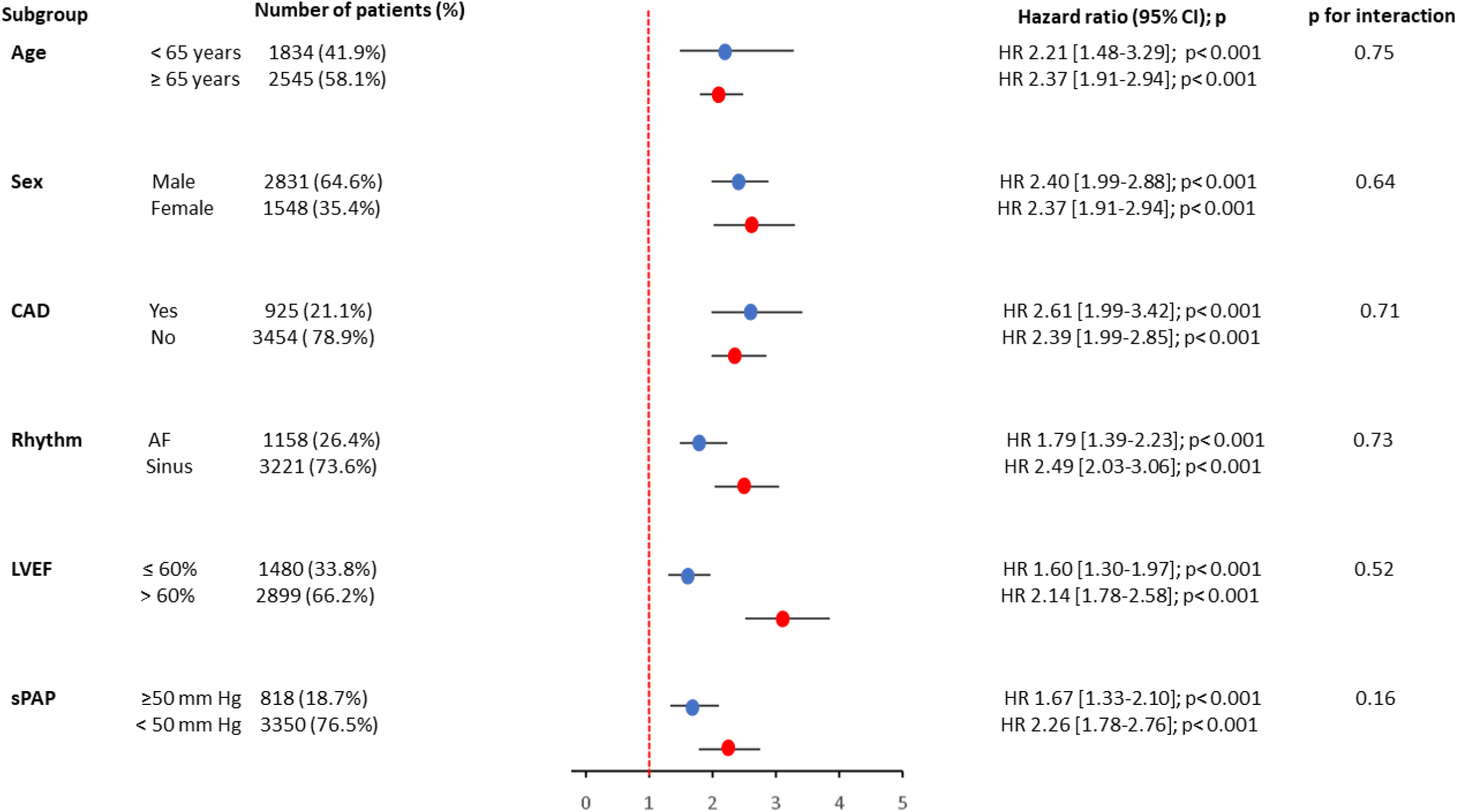
**Subgroup analysis** Forrest plot for the risk of all-cause mortality associated with right ventricular (RV) dysfunction in subgroups of patients with DMR. *CAD: coronary artery disease; CI: confidence interval; LVEF: left ventricular ejection fraction; sPAP: systolic pulmonary artery pressure*.

#### Outcome under medical management

Under medical management, 670 deaths (36.7% of non-operated patients) were recorded. Ten-year estimated survival under medical management was 42±4% for patients with RVD *vs*. 58±2% for patients with normal RV function (Log Rank *p<0.001*) (***Figure 3A***). On Cox multivariable analysis, after adjustment for age, sex, presence of symptoms, AF, LVESD ≥ 40mm, LVEF≤ 60%, LAVI ≥ 60 ml/m², sPAP≥ 50 mm Hg and MR severity, RVD remained independently associated with increased mortality (adjusted HR [95%CI] =1.39 [1.12-1.72]; *p=0.003*) (***Figure 3B***).

**Figure 3:**
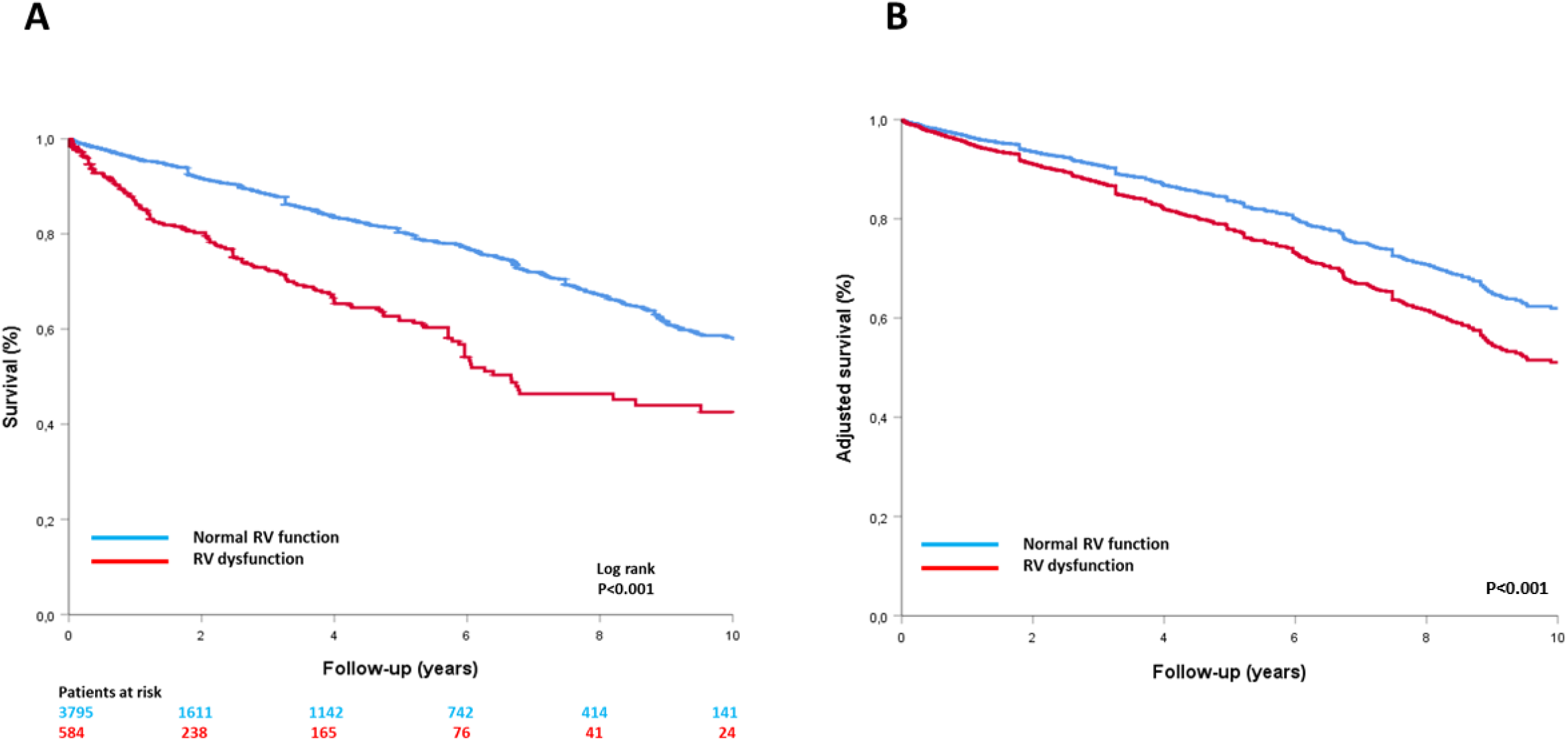
Survival curves under medical management. Kaplan-Meier (A) and Cox adjusted (B) 10-year survival curves according to right ventricular function in patients under conservative management. *RV: right ventricular*.

#### Post–Mitral Valve Surgery Outcome

Mitral valve surgery was performed in 2552 patients (57.6%) of which, 370 (14.5%) died during follow-up. One-month post-operative mortality was of 2.2% in patients with normal RV function *vs.* 5.0% in patients with RVD (*p<0.001*). On multivariable logistic regression, RVD remained associated with decreased 1-month post-operative survival after adjustment for age, sPAP, EuroSCORE II and time from baseline echocardiography to surgery (adjusted OR [95%CI] =1.84 [1.13-2.99]; *p=0.014*).

Ten-year estimated post-operative survival was 85±4% for patients with RVD vs. 91±1% for patients with normal RV function (Log Rank *p<0.001*) (***Figure 4A***). On Cox multivariable analysis, RVD remained independently associated with reduced long-term post-operative survival (adjusted HR [95%CI] =1.56 [1.08-2.27]; *p=0.019*) (***Figure 4B***).

**Figure 4:**
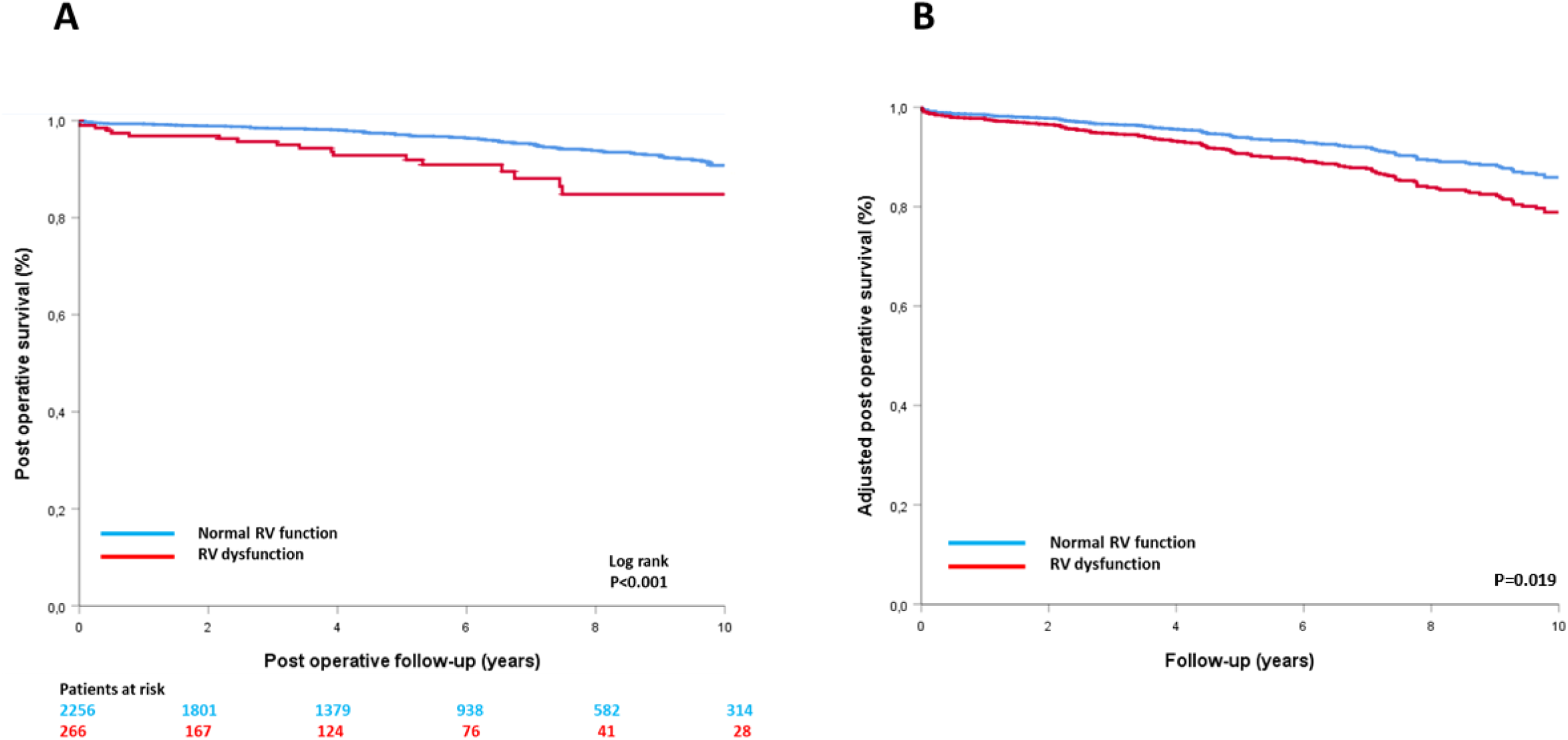
Post-operativen survival curves. Kaplan-Meier (A) and Cox adjusted (B) 10-year post operative survival curves according to right ventricular function. *RV: right ventricular*.

### Impact of mitral valve surgery in patients with RV dysfunction

Among the 584 patients with RVD, 266 (45.5%) underwent mitral valve surgery during follow-up, including 179 within 3 months of diagnosis. Patients who underwent surgery within 3 months of diagnosis were younger and had a lower EuroSCORE II (both *p<0.001*). They were more symptomatic with more severe MR, more likely to have a flail leaflet, and had lower LVEF than patients not operated on within 3 months of baseline echocardiography (all *p <0.001)* (**Table 3**).

**Table 3:** Baseline characteristics of patients with right ventricular dysfunction according to initial management.

| Characteristics | Surgery ≤ 3 months | Medical management | P-Value |
| --- | --- | --- | --- |

|  | n=179 | or surgery > 3months<br>n=405 |  |
| --- | --- | --- | --- |
| **<u>Clinical characteristics</u>** |  |  |  |
| Age, years | 70 ± 11 | 76 ± 12 | <**0.001** |
| Female gender (% , n) | 31.3 (56) | 35.1 (141) | 0.39 |
| Body mass index, kg/m<sup>2</sup> | 26.4 ± 4.2 | 25.2 ± 4.9 | **0.004** |
| Hypertension (% , n) | 31.3 (56) | 33.8 (137) | 0.57 |
| Diabetes mellitus (% , n) | 10.1 (18) | 13.1 (53) | 0.34 |
| Coronary artery disease (% , n) | 30.7 (55) | 24.4 (99) | 0.127 |
| EuroSCORE II, % | 1.4 ± 0.8 | 2.1 ± 1.6 | <**0.001** |
| Symptoms (% , n) | 78.1 (139) | 62.2 (247) | <**0.001** |
| Atrial fibrillation (% , n) | 37.4 (67) | 43.7 (177) | 0.18 |
| **<u>Echocardiographic characteristics</u>** |  |  |  |
| LV end-diastolic diameter, mm | 57 ± 7 | 55 ± 8 | **0.029** |
| Indexed LV end-diastolic diameter, mm/m<sup>2</sup> | 31 ± 5 | 30 ± 5 | 0.51 |
| LV end-systolic diameter, mm | 37 ± 8 | 37 ± 8 | 0.96 |
| Indexed LV end-systolic diameter, mm/m<sup>2</sup> | 20 ± 4 | 20 ± 5 | 0.12 |
| LV ejection fraction, % | 61 ± 10 | 57 ± 11 | <**0.001** |
| LA volume index, mL/m<sup>2</sup> | 76 ± 29 | 70 ± 33 | 0.06 |
| EROA, mm<sup>2</sup> | 53 ± 22 | 40 ± 23 | <**0.001** |
| MR regurgitant volume, ml | 77 ± 31 | 60 ± 33 | <**0.001** |
| Bileaflet prolapse, (% , n) | 38.0 (68) | 57.8 (234) | <**0.001** |
| Posterior prolapse, (% , n) | 51.4 (93) | 69.9 (283) | **0.004** |
| Flail Leaflet, (% , n) | 47.2 (75) | 23.9 (87) | <**0.001** |
| PA systolic pressure, mmHg | 54 ± 19 | 51 ± 17 | 0.065 |

Continuous variables are expressed as mean ±1 standard deviation and categorical variables are expressed as percentages and numbers.
Abbreviations: COPD: chronic obstructive pulmonary disease; EROA: effective regurgitant orifice; LA: left atrial; LV: left ventricular; NYHA= New York Heart Association; PA: pulmonary artery.

Ten-year estimated survival was 73±4% for patients with RVD who underwent surgery within 3 months of diagnosis *vs.* only 38±4% for patients with RVD not operated within 3 months (Log Rank *p<0.001*) (***Figure 5A***). On Cox multivariable analysis, age ≥ 65 years (adjusted HR [95%CI] =5.08 [2.73-8.47]; *p<0.001*) and sPAP ≥ 50 mm Hg (adjusted HR [95%CI] =1.70 [1.23-2.35]; *p=0.001)* were independently associated with mortality whereas surgery ≤ 3 months was associated with a better outcome (adjusted HR [95%CI] =0.53 [0.35-0.81]; *p=0.003*) in patients with RVD (***Figure 5B***).

**Figure 5:**
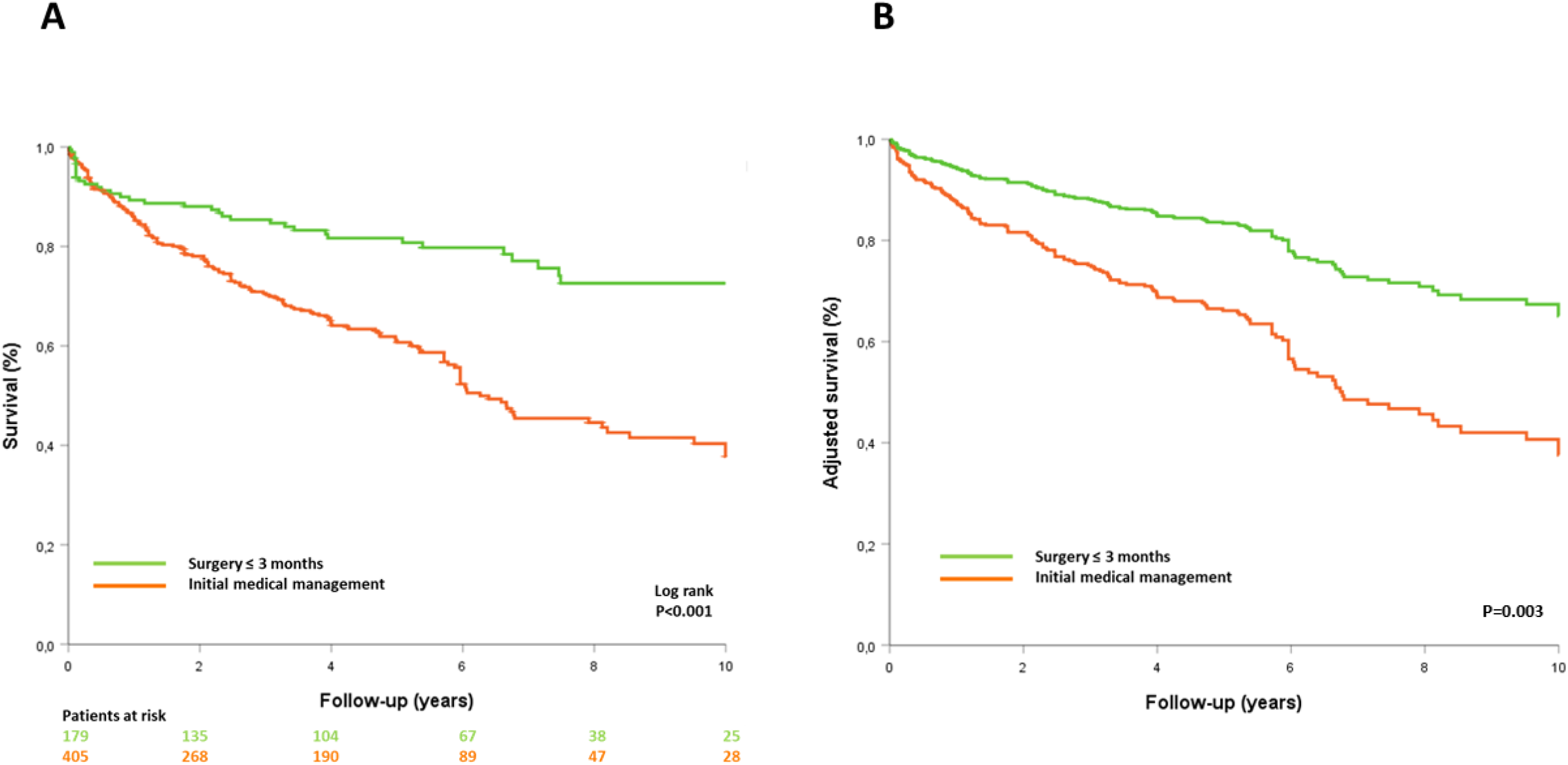
**Survival curves according to management in RVD** Kaplan-Meier (A) and Cox adjusted (B) 10-year survival curves in patients with right ventricular dysfunction according to initial management.

## DISCUSSION

The present study, based on a multicenter large cohort of patients with significant DMR managed in routine clinical practice, demonstrates that RVD assessed by transthoracic echocardiography is a major and independent determinant of long-term survival in response to conservative or surgical management. Indeed, RVD (observed in 13.3% of patients) has a strong impact on mortality, persisting after adjustment for covariates known to be major determinants of outcome in DMR, such as age, comorbidity, AF, LV dilatation and dysfunction, left atrial volume and pulmonary pressures. Importantly, the current results demonstrate that RVD is not simply predictive of greater mortality under medical management but is also associated with worse survival after MR surgical correction, despite the substantial mortality reduction associated with surgery. Therefore, mitral valve surgery should be discussed before the onset of RVD.

### Prevalence of RV dysfunction

RV function has not been extensively studied in MR and only small series have been published so far (9,23,24), complicating the estimation of the prevalence of RVD in patients with significant DMR. Recent data on transcatheter mitral valve repair report a high prevalence of RVD assessed by echocardiography in primary MR, ranging from 30% to 39% of patients, but this is a selected population of older patients with severe symptomatic MR.(25) Le Tourneau et al (11) also reported a high RVD prevalence of 30% in a population of 208 patients undergoing mitral valve surgery for organic MR. However, in this study (11), RV function was assessed by radionuclide angiography with poor correlation between RV ejection fraction and RV S-wave velocity measured by echocardiography. Thus, the reported prevalence of RVD in DMR depends on the method employed for its identification and on the studied population. No large study has yet reported the prevalence of RVD assessed by routine clinical echocardiography in a population of consecutive patients with DMR. We identified RVD in 13.3% of patients with significant DMR. Interestingly, LVEF was reported as normal in half of these patients and RVD was identified in all patient subgroups illustrating that it should be routinely investigated.

#### Mechanisms and determinants of RV dysfunction in DMR

The thin right ventricular wall is very sensitive to changes in loading conditions, especially for pressure overload explaining RV systolic performance alteration in case of primary(3) or secondary pulmonary hypertension.(1,2) In MR, the increase in left atrial pressure results in a backward elevation of pulmonary venous pressure, pulmonary capillary wedge and sPAP. In addition, pulmonary vascular remodeling with abnormal vasoconstriction contributes to the rise in pulmonary pressures.(11,26) Nevertheless, if there is a clear relationship between sPAP and RV function, this relationship is moderate for Le Tourneau et al. (11) and sPAP is not the only factor associated with RVD.(11,27) Indeed, as identified in the present study, increased sPAP was the factor most strongly associated with RVD but other parameters including age, MR severity, LAVI and LV systolic dysfunction were also independently associated with RVD in patients with DMR. In addition, RVD predicted mortality after adjustment to pulmonary hypertension and was also associated with poor survival in patients with sPAP< 50 mmHg.

Severe MR leads to LV remodeling to compensate for chronic volume overload by eccentric hypertrophy, resulting in a more spherical shape of the LV. This remodeling will alter the RV performance through a double mechanism: (i) the interventricular septum will curve, resulting in altered RV free wall contraction with increasing ventricular interdependence while (ii) the spherical shape of the LV will also generate increased intrapericardial constraint, transmitted through ventricular interdependence into the RV. (28,29) It is also important to point out that in severe MR, LV dysfunction is often masked by an apparently preserved LVEF but this contractile dysfunction will affect RV systolic function, notably through the interventricular septum. Importantly, a LVEF≤ 60% is found in 54% of patients with altered RV ejection fraction, suggesting that RVD may reflect chronic LV systolic impairment which is the consequence of long-standing MR. (11,28) Accordingly, in our study, 50,7% of patients with RVD had a LVEF≤60%.

#### Outcome of RV dysfunction in DMR

Over the past few years, the prognostic importance of RVD has been increasingly emphasized in many cardiovascular diseases.(1–5) Nevertheless, regarding DMR, data on RVD are scarce. Doldi et al.(25) recently reported that RVD assessed by echocardiography was associated with a reduced survival in patients undergoing transcatheter mitral valve repair for primary MR. Older studies also reported that patients with DMR and LV dysfunction undergoing mitral valve surgery had reduced survival in patients with associated RVD. (11,30,31) However, the present study is the first to report the prognostic impact of RVD assessed by echocardiography in clinical routine in a population of consecutive patients with significant DMR. The current study shows that RVD has a strong impact on mortality, partially attenuated but persisting after adjustment for covariates known to be major determinants of outcome in DMR including LV dysfunction, AF and pulmonary hypertension, under medical or surgical management. The adverse effect of RVD was consistently observed in subgroups of patients with MR even in patients without apparent LV dysfunction or pulmonary hypertension suggesting that RV function should be systematically assessed in case of significant primary MR.

Although mitral valve surgery was associated with reduced mortality in patients with RVD compared to conservative management, excess mortality persisted after surgery in these patients, and 1-month postoperative mortality was more than 2-fold higher than in patients without RVD. Therefore, surgery should be considered before impairment of RV systolic performance occurs. Indeed, while early surgery is often considered as the ideal treatment for patients with severe DMR, it is rarely performed, especially in elderly patients with commodities, and we need to identify all the potential risk markers in order to reduce under-treatment of this disease. Finally, our results strongly suggest that assessment of RV function by echocardiography should be performed systematically for risk stratification of patients with DMR.

### Limitations

The current study is subject to the limitations inherent to observational registries with retrospective follow-up data. However, echocardiographic data were collected prospectively at each center by multiple operators, which allowed us to constitute the largest international cohort of patients with DMR quantified in clinical routine. Transthoracic echocardiography is limited in the assessment of RV function, due to the asymmetrical and crescentic shape of the RV. Nevertheless, it remains the first-line imaging technique and the cornerstone for diagnosis of RVD in clinical practice. While all centers applied the guidelines available for the comprehensive determination of RV function based on qualitative and quantitative data, the databases available at time of patient diagnosis in most cases did not have fields available for recording all quantitative measures that form part of the basis of the classification as RV dysfunction. Thus, the TAPSE was the most frequently stored quantitative variable recorded (n=2186). When the population was stratified according to stored TAPSE≥or < 17 mm, the results in term of outcome was not different from the categorical RV function classification. Thus, the quantitative or integrative assessment of RV function are linked to outcome in a very similar manner and do not affect the results of the study, underscoring for the first time the importance of RV dysfunction, incrementally to pulmonary hypertension and all baseline characteristics, in defining the prognosis of patients with DMR. Assessment of RV function by echocardiography can be challenging and cardiac magnetic resonance, which appears to play an interesting role in the risk stratification of patients with mitral valve prolapse (32), should include assessment of RV volumes and function when performed in this indication. Data regarding the cause of death and occurrence of heart failure were not available, which precluded these analyses. Finally, our results are only applicable to patients with DMR due to mitral valve prolapse, and further studies are needed for other subsets of patients with organic MR.

## CONCLUSION

This study is the first to demonstrate in a large cohort that RVD assessed by echocardiography is a powerful predictor of survival in patients with significant DMR due to mitral valve prolapse. Although patients with RVD (13.3%) exhibited increased mortality under medical and surgical management, they should still be considered for surgery to improve their outcomes. To avoid the excess mortality observed after mitral valve surgery related to RVD, surgery should be discussed before RV dysfunction occurs.

## Data Availability

The data that support the findings of this study are available from the corresponding author on reasonable request.

## Acknowledgments

none

